# Real-life course and effectiveness of melatonin treatment for sleep disturbances in children with autism spectrum disorder

**DOI:** 10.1101/2023.06.13.23291321

**Authors:** Hadar Sadeh, Gal Meiri, Dikla Zigdon, Michal Ilan, Michal Faroy, Analya Michaelovski, Yair Tsadaka, Ilan Dinstein, Idan Menashe

## Abstract

**Objective:** Melatonin is considered the most effective pharmacological treatment for the sleep disturbances that are reported in >50% of children with autism spectrum disorder (ASD). However, real-life data about the long-term course and effectiveness of melatonin treatment in children with ASD is lacking.

**Methods:** In this retrospective cohort study, we assessed the adherence to melatonin treatment and its effect on sleep quality and daytime behavior in children with ASD via a parental phone questionnaire of children in the Azrieli National Center for Autism and Neurodevelopment Research (ANCAN) database. Cox regression analysis was used to assess the effect of key demographic and clinical characteristics on treatment adherence.

**Results:** Melatonin was recommended for ∼8% of children in the ANCAN database. These children were characterized by more severe autistic symptomatology. The median adherence time for melatonin treatment exceeded 88 months, with the most common reason for discontinuation being a lack of effectiveness (14%). Mild side-effects were reported in 14% of children, and 86%, 54%, and 45% experienced improvements in sleep onset, sleep duration and night awakenings, respectively. Notably, melatonin also improved the daytime behaviors of >28% of the children. Adherence to treatment was independently associated with improvements in night awakenings and educational functioning (aHR=0.142, 95%CI=0.036-0.565; and aHR=0.195, 95%CI=0.047-0.806, respectively).

**Conclusions:** Our findings highlight the real-life safety and effectiveness of melatonin treatment in children with ASD. Increasing the awareness of this treatment among families and health providers will help to reduce the burden associated with sleep disturbances in children with ASD and their families.

## Introduction

Sleep disturbances are highly prevalent in children with autism spectrum disorder (ASD), with an estimated prevalence ranging between 30–90%.^1–4^ These sleep disturbances, which may occur from early life into adolescence and adulthood,^5–7^ include, but are not limited to, delayed sleep onset, multiple night awakenings, early morning awakenings, low sleep efficiency, delayed circadian phases, and shortened total sleep time.^4, 8, 9^ Children with ASD who experience sleep disturbances usually manifest more severe core symptoms of ASD, such as poorer social capabilities, more restricted- repetitive behaviors, and more communication and language problems than their non-ASD counterparts.^10–12^ Moreover, sleep problems in children with ASD are also associated with the manifestation of other behavioral problems, such as irritability, aggression, anxiety, affective problems, hyperactivity, attentional deficits and poorer functional and adaptive skills.^6, 7, 13, 14^

Melatonin is an endogenous hormone produced in the pineal gland.^15^ Its secretion is regulated through light-dark cues, by the biologic clock, the supra-chiasmatic nucleus (SCN) in the hypothalamus. It plays a role in various physiologic functions, including sleep promotion and circadian modulation through feedback activity on the SCN and peripheral tissues.^15^ Due to its key function in the circadian rhythm, melatonin is used to treat various forms of sleep problems,^16^ including those manifested by children with ASD.^17, 18^

Melatonin is usually prescribed for treating sleep latency in children with ASD, but it has also been reported to increase total sleep duration and sleep efficiency in those children^19, 20^ and to reduce night awakenings (particularly for the prolonged-release formulation).^21, 22^ Moreover, melatonin treatment before bedtime has been found to improve daytime behavior and core symptoms of children with ASD^20, 23, 24^. Importantly, melatonin is considered to be safe, with only a few transient or mild reported side effects, such as early morning awakenings, morning drowsiness, headache, mood swings and irritability.^19–21, 25^

Despite the compelling evidence regarding the positive effect of melatonin treatment on sleep disturbances in children with ASD, only 7–13%^17, 20^ of children with ASD are prescribed melatonin for sleep problems. In addition, real-life and long-term data on the course and effectiveness of melatonin treatment are sparse. Finally, the clinical characteristics associated with adherence to melatonin treatment are vague. For these reasons, we studied the real-life prevalence, course, and effectiveness of melatonin treatment in a representative sample of children with ASD in Israel.

## Methods

### Study design

We conducted a retrospective cohort study of children diagnosed with ASD who are registered in the database of the Azrieli National Center for Autism and Neurodevelopment Research (ANCAN), Israel.^26^ Inclusion and exclusion criteria for the study sample are shown in **Figure S1**, available online. We included children whose database records contained an indicator or recommendation for melatonin use and whose parents signed an informed consent form allowing us to contact them for research purposes. We contacted the parents of those children by phone and asked them to answer a brief questionnaire about their experience with the melatonin treatment. We excluded children without contact details (e.g., phone number or email address), those whose parents refused to participate in the study, and those whose parents indicated that the children had not been given melatonin or had taken it for less than a week.

### Data collection

Data about melatonin use, effectiveness and side effects were obtained from the above-described parental phone questionnaires. The questionnaire (which is presented in **Supplementary file 1**, available online) comprised two parts: 1) eight questions, adopted from other questionnaires,^27, 28^ that include details about melatonin dosages, usage patterns (time and frequency), duration of treatment, reason for stopping or continuing treatment, side effects, and source of the medicine; and 2) eight questions that were designed to assess the child’s sleep and general daily functioning and behavior, on a scale of 1 to 5 (1 – major negative effect; 2 – minor negative effect; 3 – no effect; 4 – minor positive effect; and 5 – major positive effect), as described previously.^27–29^

Sociodemographic, behavioral, and clinical data about participating children were obtained from the ANCAN database. Clinical variables included the Autism Diagnostic Observation Schedule™, Second Edition (ADOS®-2)^30^ comparison score, the DSM-5 ASD severity levels, the preschool language scale, fourth edition (PLS4) total score,^31^ a cognitive score based on either the Bayley Scales of Infant and Toddler Development-third edition (Bayley-III)^32^ or the Wechsler Preschool and Primary Scale of Intelligence—version three (WPPSI-III),^33^ The General Adaptive Composite (GAC) score of the Adaptive Behavior Assessment System-II (ABAS-II)^34^, and data about the child’s sleep quality based on the Children’s Sleep Habits Questionnaire (CSHQ).^35^ Additional data about chronic comorbidities and medication use were obtained from the children’s medical files.

### Statistical analysis

Standard univariate statistics were used to assess differences in sociodemographic and clinical characteristics between the study cohort (i.e., children who were reported to use short-release melatonin or were recommended to use it) and other children in the ANCAN database as well as between children whose parents reported melatonin had/did not have an effect on their sleep. Kohen’s kappa was used to assess the concordance between different effects of melatonin treatment. Kaplan-Meir and Cox regression analyses were used to assess sociodemographic and clinical variables associated with adherence to melatonin treatment. Statistical significance in all these analyses was determined at a p-value of <0.05. All analyses were carried out using SPSS Software Version 28.

## Results

Among the 1,355 children with ASD in the ANCAN database (November 2021), there an indication for melatonin recommendation or use for only 107 (8%) children (**Figure S1**, available online). A comparison of the sociodemographic and clinical characteristics between children with ASD who were treated with melatonin and those who were not is presented in **Table 1**. Children who were given melatonin were diagnosed, on average, six months earlier than children who were not (35.1±15.3 months vs. 41.4±19.3 respectively; p-value = 0.002). Children who were treated with melatonin had more severe ASD symptoms, according to both DSM-5 criteria and the ADOS-2 comparison score, and they also had lower cognitive and adaptive behavior abilities (**Table 1**). As expected, children who were treated with melatonin had more severe sleep disturbances, according to the CSHQ total and subscales scores, compared to their counterparts who were not.

**Table 1:** Comparison of Sociodemographic and Clinical Characteristics between Children with ASD with and without a Melatonin Recommendation.

| Variable |  | Melatonin recommended<br>(n=107) | Melatonin not recommended<br>(n=1248) | P-value |
| --- | --- | --- | --- | --- |
| Age at diagnosis in months (mean±SD) |  | 35.1 (+-15.3) | 41.4 (+-19.3) | **0.002<sup>a</sup>** |
| Sex (% male) |  | 88 (82%) | 989 (79%) | 0.561<sup>c</sup> |
| Ethnicity (% Jewish) |  | 89 (84%) | 1001 (82%) | 0.705<sup>c</sup> |
| ADOS-2 comparison score (mean±SD) |  | 7.7 (+-2.2) | 6.9 (+-2.2) | **0.002<sup>a</sup>** |
| DSM-5 severity level (A criteria) | Requiring support | 10 (10%) | 234 (21%) | **0.004<sup>c</sup>** |
|  | Requiring substantial support | 42 (41%) | 496 (44%) |  |
|  | Requiring very substantial support | 50 (49%) | 391 (35%) |  |
| DSM-5 severity level (B criteria) | Requiring support | 14 (13%) | 290 (23%) | **<0.001<sup>c</sup>** |
|  | Requiring substantial support | 42 (39%) | 575 (46%) |  |
|  | Requiring very substantial support | 45 (42%) | 263 (21%) |  |
| Cognitive score (mean±SD) |  | 71.6 (+-18.6) | 76.1 (+-19.9) | 0.067<sup>a</sup> |
| ABAS GAC score (mean±SD) |  | 57.3 (+-9.5) | 67.3 (+-15.9) | **<0.001<sup>a</sup>** |
| PLS score (mean±SD) |  | 63.8 (+-24) | 62.5 (+-32) | 0.780<sup>a</sup> |
| CSHQ | Total sleep score (mean±SD) | 51.8 (+-10.3) | 46.1 (+-9.3) | **<0.001<sup>a</sup>** |
|  | Sleep onset delay (mean±SD) | 2.3 (+-0.8) | 1.8 (+-0.8) | **<0.001<sup>a</sup>** |
|  | Night wakings (Mean±SD) | 5.2 (+-1.8) | 4.7 (+-1.7) | **0.026<sup>a</sup>** |
|  | Sleep duration (mean±SD) | 5.1 (+-1.8) | 4 (+-1.5) | **<0.001<sup>a</sup>** |

Note: <sup>a</sup> T- test; <sup>b</sup> Fisher's exact test; <sup>c</sup> Chi-square test (1 df)

### Characteristics of the melatonin-treatment cohort

Overall, the parents of 78 children (73%) for whom there was an indication for melatonin recommendation or use completed our phone questionnaire and reported adherence to melatonin treatment for at least one week. Notably, the median adherence time for those children exceeded 88 months (**Figure 1**), with only 26 children (33.5%) discontinuing the treatment for various reasons, including lack of effectiveness (14.1%), natural improvement (8.9%), loss of effectiveness (7.7%), side effects (3.8%), starting other medications (3.8%), child’s refusal (2.6%), doctor’s recommendation (2.6%), medication cost (1.3%), or starting a behavioral sleep intervention (1.3%) (**Table S1**, available online).

**Figure 1.**
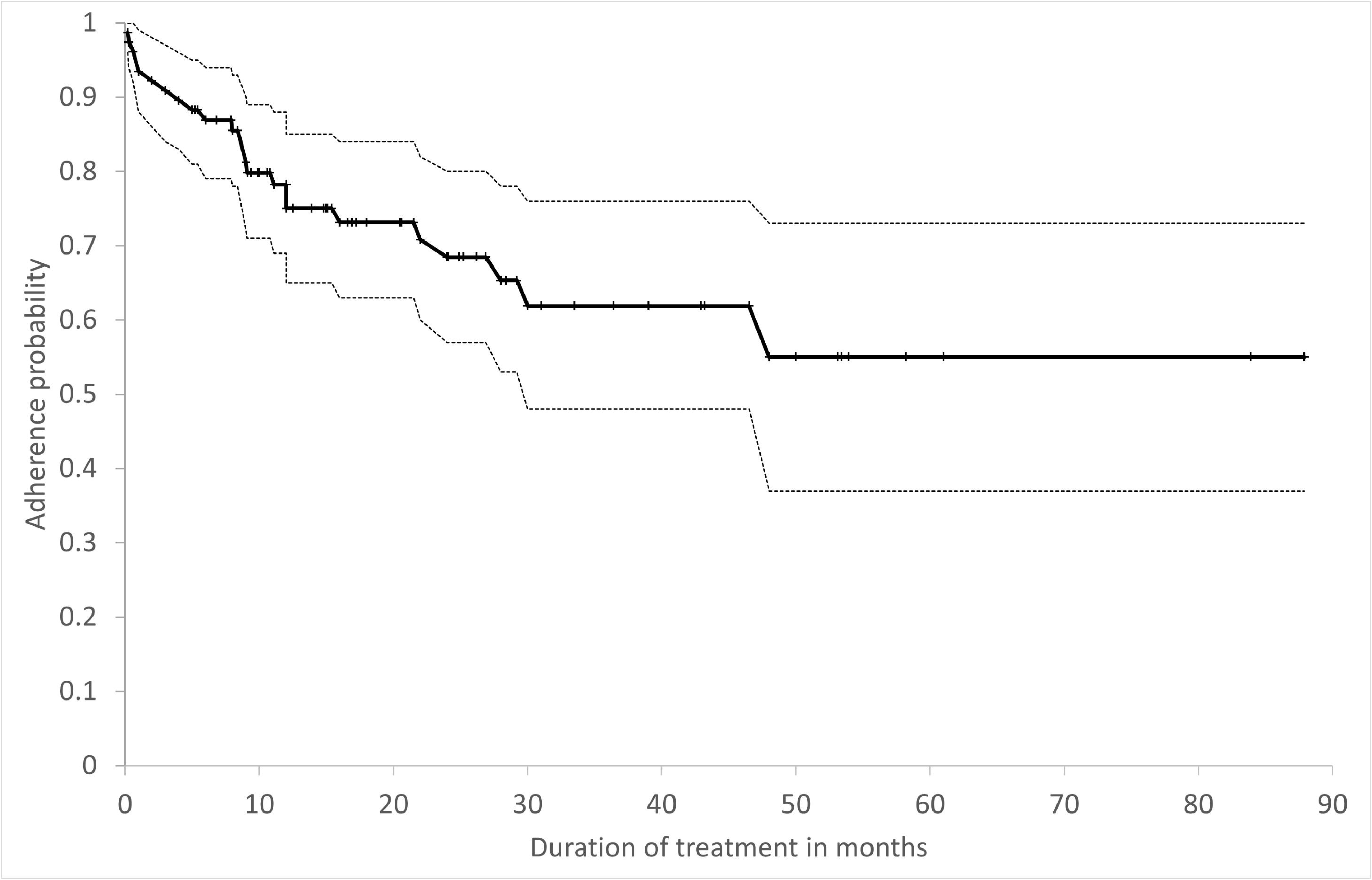
Kaplan-Meier Curve of Melatonin Treatment Adherence over Time. The cumulative adherence probability to melatonin treatment over time is depicted by a continuous black line with 95% confidence intervals in black dashed lines.

The sociodemographic and clinical characteristics of the children who were treated with melatonin, and the association of these characteristics with treatment discontinuation, are presented in **Table 2**. None of these characteristics was significantly associated with treatment discontinuation, although the use of other psychoactive drugs concurrently with melatonin treatment was marginally associated with a lower probability of treatment discontinuation (HR = 0.452, 95%CI = 0.194–1.052, respectively). Characteristics associated with melatonin treatment and their effect on treatment discontinuation are also presented in **Table 2**. The age at treatment initiation ranged between 1.6 to 9.6 years, and the mean final melatonin dosage was 3.9 (±2.7) mg. Most of the parents (77%) gave the treatment every day before bedtime, while the others gave melatonin only on school days, or when the child had serious sleep onset difficulties. Interestingly, the parents of 14 children (18%) purchased an imported over-the-counter (OTC) formulation of melatonin, without prescription. The parents of 11 children (14%) reported mild side effects of melatonin treatment, including crying, irritability/hyperactivity, morning drowsiness, rash, vomiting, fever, increased appetite, and abdominal pain (**Table S1**, available online). None of the treatment characteristics or its side effects was significantly associated with treatment discontinuation.

**Table 2:** Characteristics Associated with Discontinuation of Melatonin Treatment.

| Variables |  | All | HR for treatment discontinuation | 95% CI | P-value |
| --- | --- | --- | --- | --- | --- |
| **Sociodemographic and clinical characteristics** |  |  |  |  |  |
| Sex, n (%) males |  | 67 (86%) | 1.165 | 0.397-3.413 | **0.781** |
| Ethnicity, n (%) Jewish |  | 62 (79%) | 0.864 | 0.294-2.544 | **0.791** |
| Age at diagnosis in months (mean±SD) |  | 34.3+-15.6 | 0.875 | 0.618-1.239 | **0.452** |
| ADOS-2 comparison score (mean±SD) |  | 7.4+-2.2 | 1.071 | 0.875-1.312 | **0.505** |
| DSM-5 severity level (A criteria), n (%) | Requiring support | 0 (0%) | 0.539 | 0.238-1.218 | **0.137** |
|  | Requiring substantial support | 36 (46%) |  |  |  |
|  | Requiring very substantial support | 39 (50%) |  |  |  |
| DSM-5 severity level (B criteria), n (%) | Requiring support | 4 (5%) | 1.060 | 0.457-2.458 | **0.892** |
|  | Requiring substantial support | 35 (45%) |  |  |  |
|  | Requiring very substantial support | 36 (46%) |  |  |  |
| PLS score (mean±SD) |  | 63.2+-24.3 | 0.995 | 0.975-1.015 | **0.642** |
| Cognitive score (mean±SD) |  | 71.9+-18 | 1.020 | 0.993-1.047 | **0.150** |
| ABAS GAC score (mean±SD) |  | 57.3 +-10.3 | 0.992 | 0.929-1.060 | **0.820** |
| Any medical comorbidity, n (%) |  | 38 (49%) | 0.669 | 0.293-1.530 | **0.341** |
| Any neuro-developmental comorbidity,<sup>a</sup> n (%) |  | 9 (11%) | 1.162 | 0.345-3.909 | **0.809** |
| Any emotional-behavioral comorbidity,<sup>b</sup> n (%) |  | 7 (9%) | 0.038 | 0.000-6.811 | **0.217** |
| ADHD, n (%) |  | 25 (32%) | 0.605 | 0.245-1.494 | **0.276** |
| Any other psychoactive medications,<sup>c</sup> n (%) |  | 37 (47%) | 0.452 | 0.194-1.052 | **0.066** |
| Antipsychotic medications, n (%) |  | 19 (24%) | 0.729 | 0.272-1.955 | **0.530** |
| Stimulant medications, n (%) |  | 15 (19%) | 0.796 | 0.291-2.178 | **0.657** |
| **Treatment characteristics** |  |  |  |  |  |
| Age at treatment beginning in months (mean±SD) |  | 51.9 (+-20) | 1.118 | 0.890-1.404 | **0.336** |
| Final dosage in mg (mean±SD) |  | 3.98 (+-2.7) | 0.920 | 0.769-1.102 | **0.367** |
| Treatment course | Times per week (mean±SD) | 6.3 (+-1.5) | 1.385 | 0.843-2.276 | **0.199** |
|  | Continuity (no cessation of more than several days), n (%) | 63 (81%) | 1.798 | 0.535-6.040 | **0.342** |
| Source of medication – imported, as dietary supplement, n (%) |  | 14 (18%) | 0.490 | 0.145-1.656 | **0.251** |
| Any side effects, n (%) |  | 11 (14%) | 1.919 | 0.710-5.188 | **0.199** |

*Note: <sup>a</sup> including- epilepsy, Down's syndrome, global and motor developmental delay, hydrocephalus; <sup>b</sup> including- anxiety, enuresis, encompresis, oppositional-defiant disorder; <sup>c</sup> including- Prothiazine, anti-psychotics, anti-convulsants, SSRI, stimulants, cannabis*

**Table 3:** Characteristics of Treatment Effectiveness and its Effect on Adherence.

| **Reported effect <sup>a</sup>** | **N (%)** | **Univariate HR for treatment cessation** | **95% CI** | **p value** | **Multivariate HR for treatment cessation** | **95% CI** | **p-value** |
| --- | --- | --- | --- | --- | --- | --- | --- |
| **Effect on sleep quality** |  |  |  |  |  |  |  |
| Sleep onset | 67 (86%) | 0.535 | 0.199-1.435 | 0.214 | 1.055 | 0.333-3.343 | **0.928** |
| Sleep duration | 42 (54%) | **0.376** | **0.161-0.878** | **0.024** | 1.230 | 0.411-3.686 | **0.711** |
| Night awakening | 35 (45%) | **0.208** | **0.071-0.610** | **0.004** | **0.142** | **0.036-0.565** | **0.006** |
| **Effect on daily behavior** |  |  |  |  |  |  |  |
| Educational functioning | 22 (28%) | **0.271** | **0.080-0.913** | **0.035** | **0.195** | **0.047-0.806** | **0.024** |
| Mood | 16 (21%) | 0.468 | 0.139-1.573 | 0.220 | 0.191 | 0.005-7.344 | **0.374** |
| Temper tantrums | 8 (10%) | 0.290 | 0.039-2.152 | 0.226 | 2.212 | 0.058-83.932 | **0.669** |
| Communication | 7 (9%) | 0.412 | 0.056-3.050 | 0.385 | 2.764 | 0.249-30.628 | **0.407** |
| Sensory regulation | 5 (6%) | 1.093 | 0.256-4.655 | 0.905 | 4.119 | 0.119-142.282 | **0.434** |

*Note: <sup>a</sup> at least some effect*

### Effect of melatonin treatment

The parents of 70 children (90%) reported that melatonin improved their children’s sleep, with 86%, 53%, and 45% reporting that melatonin treatment influenced the sleep onset, sleep duration, and night awakenings, respectively (**Figure 2A**). In addition, the parents of 27 children (35%) reported that melatonin had an additional effect on their children’s daytime behavior, with better educational functioning, improved moods, reduction in tantrums, better communication abilities and better sensory regulation being reported in 28%, 21%, 10%, 9%, and 6% of children, respectively (**Figure 2B**). Of note, a moderate and statistically significant concordance was seen between the effect of melatonin treatment on different sleep and daytime behaviors, with the most significant associations being between sleep duration and night awakening (kappa = 0.459; p-value <0.001), and between tantrum reduction and mood (kappa = 0.571; p-value <0.001) (**Figure 2C**).

**Figure 2.**
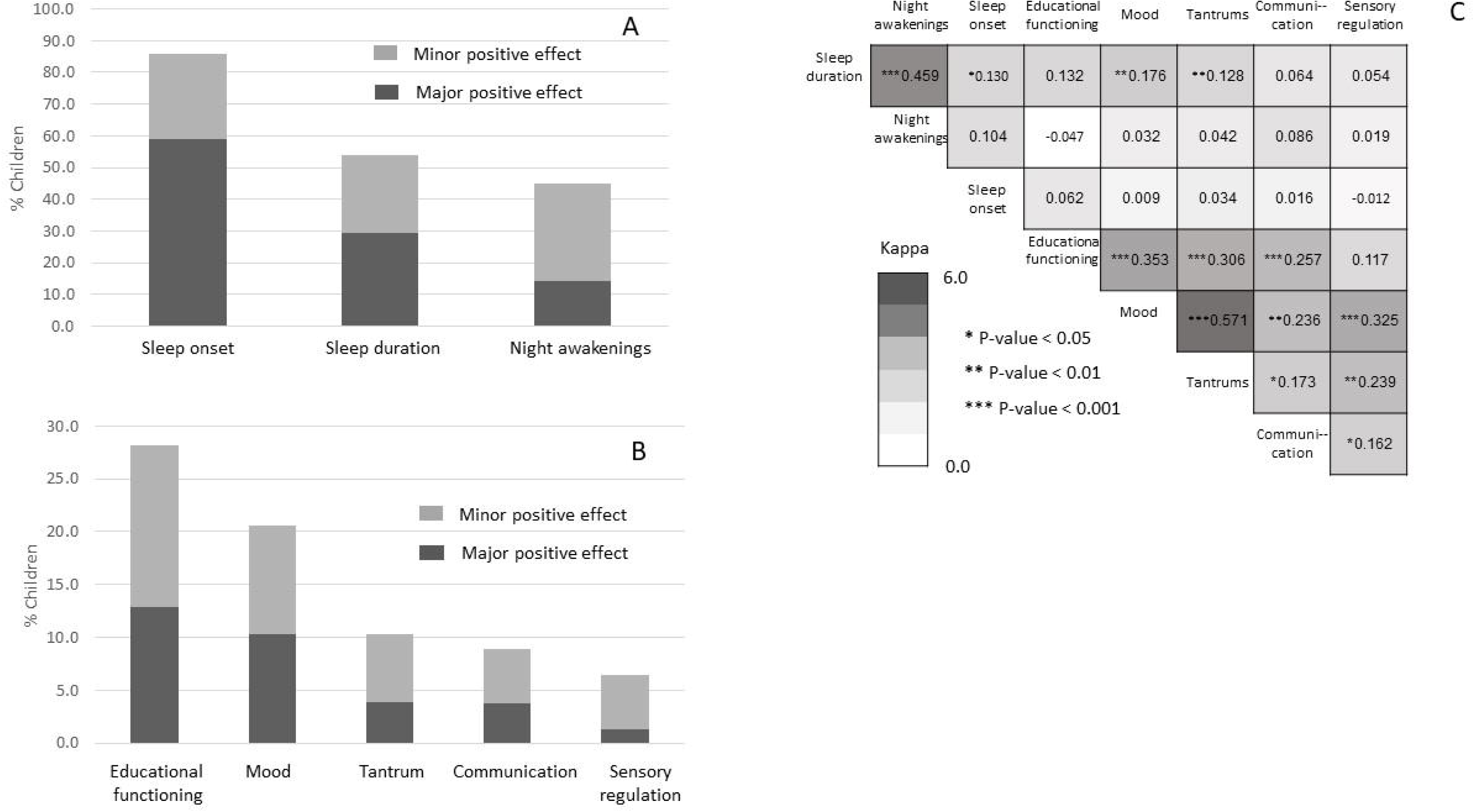
Effect of Melatonin on Sleep and Daytime Behaviors of Children with ASD. (A)&. (**B**) Rates of children whose parents reported an effect of melatonin treatment on their sleep parameters (A) and daytime behaviors (B). (**C**) Kohen’s kappa coefficients for the concordance between different effects of melatonin treatment.

Finally, both univariable and multivariable Cox regression models were used to assess the association between the reported effect of melatonin treatment on sleep and daytime behavior and treatment adherence. Improvements in nighttime awakening, sleep duration and educational functioning were significantly associated with a decreased likelihood to discontinue melatonin treatment (HR = 0.208, 95%CI = 0.071–0.610; HR = 0.376, 95%CI = 0.161–0.878; and HR = 0.271, 95%CI = 0.080–0.91, respectively) in the univariate analysis. However, because of the relationship between the effects of melatonin treatment on sleep duration and night awakenings (see **Figure 2C**), only the effects of night awakenings and educational functioning remained significantly associated with treatment adherence in the multivariate analysis (aHR = 0.142, 95%CI = 0.036-0.565; and aHR = 0.195, 95%CI = 0.047–0.806, respectively).

## Discussion

To the best of our knowledge, this is the first study to examine the real-life adherence to melatonin treatment and its effect on both sleep and daytime behaviors of children with ASD. In our sample, only 8% of ASD children were advised to take melatonin to ameliorate their sleep problems. This 8% prevalence of melatonin treatment in children with ASD is slightly higher than the 3.8% melatonin usage reported in a previous study in the same population,^36^ but is similar to the reported usage of melatonin in two meta-analyses.^29, 38^ Such a relatively low prevalence of melatonin usage stands in contrast to the >50% prevalence of sleep disturbances reported in children with ASD^1, 2, 4, 37^ and is especially surprising in light of the significant effects of melatonin on the sleep quality and daytime behavior in these children, as is evident in our study and others.^20, 22^ One possible explanation for this enigma lies in the concerns of some pediatricians about the side effects of and tolerance to melatonin, which may discourage them to recommend its use.^38, 39^ Another possible reason may stem from the known under-diagnosis by clinicians of sleep problems in children with ASD.^40–42^ Finally, it should be noted that it is not impossible that the use of imported OTC melatonin in our sample might be higher than that reported by the parents.

The children who were treated with melatonin were characterized by more severe autistic symptomatology than the other children in our cohort. This may be attributed to the known association between sleep disorders and the severity of autistic symptoms, which may motivate parents to seek treatment for their children.^7, 10, 11^ The increased severity of autistic symptoms could also explain the better adherence of the children with more severe ASD symptoms. Alternatively, the better adherence could be due to the fact these children suffer from a more severe melatonin deficiency^43–45^ and consequently derive greater benefit from the melatonin treatment.

The treatment with melatonin reported in this study was generally administered according to the accepted guidelines, which recommend administration once a day before bedtime.^18^ Furthermore, most parents also complied with the recommended dose of up to 6 mg—any further increase in the dose has been reported to be ineffective in these children.^18, 46^ Indeed, the lack of an association between final melatonin dose and treatment adherence in our cohort further supports the notion that higher doses of melatonin do not guarantee a better effect.

Approximately one fifth of the children who were given melatonin in our sample took an OTC formulation that was purchased online. Such a significant use of OTC melatonin may be attributed to the relatively easy accessibility to imported OTC melatonin and the fact that melatonin treatment was not covered by the HMOs at the time of the study. Interestingly, in terms of reported adherence and effectiveness, the OTC formulation of melatonin taken by children in the study was comparable to the prescribed melatonin, despite the remarkable variation in melatonin content in such OTC formulations^47^ and the recommendation for using prescription-only melatonin preparations.^18^ Unfortunately, we did not have information about the different types of OTC melatonin taken by the children in this study, so we could not explore differences between specific types of OTC melatonin.

Melatonin has been shown to be a relatively safe treatment, and only 14% of parents in this study reported a few mild side effects that did not affect adherence to treatment. Such minor side effects have also been reported in the literature,^19–21, 25^ although the rates of side effects were a little lower in our study, possibly because of a recall bias. Of note, despite the relatively long follow-up for most of the children in our study, some of the reported long-term side effects of melatonin treatment, such as precocious puberty,^48^ could not be observed in our sample, due to relatively young age at which treatment was initiated in most of the children.

We showed that melatonin treatment improved sleep onset in most (86%) of children, but the factors that were mostly associated with the treatment adherence were improvement in night awakenings and sleep duration. Furthermore, some parents reported that melatonin treatment also improved the daytime behaviors of their children, with an improvement in educational functioning being independently associated with treatment adherence. These findings reflect the broader effect of melatonin treatment beyond its direct effect on sleep onset. Importantly, the melatonin used in our study had a short-term effect and was therefore less effective in affecting sleep problems other than sleep onset. It is possible that other formulations of melatonin (e.g., prolonged-release formulation^49^) or other sleep medications may improve adherence to melatonin administration in these children.

The results reported in this study should be interpreted in the context of the following limitations. First, despite the large cohort of children at the ANCAN database, only 8% were treated with melatonin, a fact that significantly reduced the study sample size and the statistical power of analyses exploring the melatonin effectiveness. Second, the data on the reported effects of melatonin treatment were obtained through parental questionnaires, which may be affected by both recall and information biases. Finally, we did not have data regarding the exact formulation of melatonin used by each child and therefore we could not compare the effect of different melatonin formulations on treatment adherence, sleep quality and daytime behavior.

## Conclusions

Our results indicate that melatonin is a safe treatment, with a positive effect on both sleep quality and daytime functioning of children with ASD. Yet, in real life, only a small portion of children with ASD are prescribed – and use – this treatment. We believe that the results of this study will increase the awareness of this effective treatment among families and health providers and thus help reduce the burden associated with sleep disturbances in children with ASD and their families.

## Data Availability

All data produced in the present study are available upon reasonable request to the authors

## Acknowledgements

We would like to thank the parents who participated in this study by answering our phone questionnaire. We also thank Ms. Inez Mureinik for critical review of the manuscript.

