## supplementary materials for "Real-life course and effectiveness of melatonin treatment for sleep disturbances in children with autism spectrum disorder"

Supplementary file 1- Parents'- phone questionnaire

**Part 1- Questions about the course of treatment:**^1,2^

1. When did your child start melatonin treatment? Month-Year

2. Who recommended melatonin treatment? (specialist doctor/primary care doctor/parents initiative)

3. Is your child still using melatonin treatment? Yes/no

If the answer was no:

3a. When did your child stop using melatonin? Month-Year

3b. Why did your child stop taking melatonin? Melatonin was not effective/melatonin stopped working/natural improvement in sleep/side effects/child's refusal/ doctor's recommendation/ started another medication/ other.

4. Did your child use melatonin continuously (without cession for more than a week)? Yes/no

5. How many times a week did your child take melatonin, on average?

6. What were the initial and final dosages of melatonin treatment for your child?

7. Where did you get the treatment from? Official prescription through local pharmacy/importfrom the USA?

8. Did your child experience any side effects during melatonin treatment? If yes, which side effects and how often did they happen.

**Part 2- Questions about treatment effectiveness:**^1–3^

Please rate the effect of melatonin treatment in your child for the following aspects, on a scale of 1 to 5 as follows: 1 - major negative effect; 2 - minor negative effect; 3 - no effect; 4 - minor positive effect; and 5 - major positive effect.

9. Effect of melatonin treatment on the time it takes to your child to fall asleep

10. Effect of melatonin treatment on night awakenings of your child

11. Effect of melatonin treatment on total sleep time of your child

12. Effect of melatonin treatment on educational functioning of your child

13. Effect of melatonin treatment on mood and general emotional state of your child

14. Effect of melatonin treatment on temper tantrums/outbursts of your child

15. Effect of melatonin on verbal and non-verbal communication of your child

16. Effect of melatonin on sensory regulation of your child

**Supplementary Table S1: Reported Reasons for Melatonin Treatment Discontinuation**

| **Reason for treatment cessation** | **N (%)** |
| --- | --- |
| Any | 26 (33.5%) |
| Lack of effectiveness | 11(14%) |
| Natural improvement | 7 (9%) |
| Stopped working | 6 (7.5%) |
| Started other medications | 3 (4%) |
| Side effects | 3 (4%) |
| Child's refusal | 2 (2.5%) |
| According to doctor's recommendation | 2 (2.5%) |
| Expensive | 1 (1.5%) |
| Behavioral intervention | 1 (1.5%) |

\# note that there could be more than one reason per child

**Supplementary Table S2: Reported Side Effects**

| **Side effect** | **N (%)** |
| --- | --- |
| Any | 11 (14%) |
| Crying | 2 (2.5%) |
| Irritability/hyperactivity | 2 (2.5%) |
| Morning drowsiness | 2 (2.5%) |
| Rash | 1 (1.5%) |
| Vomiting | 1 (1.5%) |
| Fever | 1 (1.5%) |
| Increased appetite | 1 (1.5%) |
| Abdominal pain | 1 (1.5%) |

**Children with ASD in the ANCAN database (November 2021) (n=1355)**

**Melatonin recommended (n= 107, 8%)**

**Completed a phone questionnaire**

**(n=78 ,73%)**

**Excluded (n=29):**

- **No contact details in the database (n=11)**
- **Refused to answer (n=3)**
- **Did not use melatonin (n=15)**

**Supplementary Figure S1. Inclusion and Exclusion Criteria for the Study Sample**
